# The association between stigmatizing attitudes towards depression and help seeking attitudes in college students

**DOI:** 10.1101/2020.07.20.20157644

**Authors:** Virgínia da Conceição, Inês Rothes, Ricardo Gusmão

## Abstract

**Objective:** Depression stigma has been considered a significant barrier to treatment and rehabilitation. This study aimed to understand the effects of gender, previous health care use, and symptomatology on depression stigma and analyze the impact of depression stigma on help-seeking attitudes.

**Methods:** A total of 969 students with a mean age of 18.87 (SD=1.49) were included in this study and completed the Depression Stigma Scale, the Attitude Toward Seeking Professional Psychological Help, the Patient Health Questionnaire-4 questionnaire, and a socio-demographic questionnaire. We analyzed data using SPSS 24.0, with a 95% confidence interval. We performed an analysis of variance using One-Way ANOVA and analyzed possible interactions between gender and previous mental healthcare groups on depression stigma and help-seeking attitudes using a Two-Way ANOVA. T-tests were used to study differences between the gender, symptomatology groups, and previous access to mental healthcare. We also executed a hierarchical linear regression to evaluate the effects of individual characteristics on Depression Stigma and Help-seeking attitudes scores.

**Results:** Participants came from all University schools, and 64.6% were women. Stigma and help-seeking attitudes are positively affected by gender and previous access to mental healthcare services. Higher personal stigma weakened help-seeking attitudes. Depressive and anxiety symptoms influenced personal depression stigma and perceived stigma; however, we detected no direct symptomatology effect on help-seeking attitudes.

**Conclusions:** Personal depression stigma has an essential effect on help-seeking attitudes, and depressive and anxiety symptoms do not. The promotion of literacy may decrease personal depression stigma and increase professional help-seeking intentions.

## Introduction

Mental and behavioral disorders are estimated to account for 12% of the global burden of disease (World Health Organization, 2001). The number of people diagnosed with depressive disorders amounts to 4.4% of the world population, and depression ranks as the first contributor to non-fatal health burden, with 7.5% of all Years Lived with Disability (YLD) (World Health Organization, 2017).

College years are a critical developmental phase that typically coincides with emerging adulthood (Arnett, 2000), growingly conceptualized as ‘late adolescence’ (Sawyer et al., 2018). Entering a college or university mainly presents a high-risk transition due to several situational changes and upturns in life stressors, including academic and interpersonal difficulties (Uehara et al., 2010). Mental illness negatively impacts academic performance, increasing school attrition odds (Alonso et al., 2018). First-year college students have been identified as a risk group for developing depression, with reports of higher prevalence among university students than non-students for the same age group (Auerbach et al., 2018).

People affected by depressive disorders face peer rejection (Moritz & Roberts, 2016), social and institutional stigmatizing responses similar to psychosis or chronic mental illness (Corrigan, 2006), with non-recognition of mental disorders as general health illness both by health professionals and society in general (McNair et al., 2002), healthcare access constraints (Thornicroft et al., 2017), and increased difficulties in finding and maintaining a paid job (Khalema & Shankar, 2014).

Thus, reducing the stigma affecting people who have a mental illness is one of the main objectives of the Mental Health Action Plan 2013–2020 of the World Health Organization (World Health Organization, 2013). Identifying predictors of stigma is an important starting point to address stigmatizing attitudes (Griffiths et al., 2008).

Much of the recent research on depression stigma has focused on two types of social stigma: personal stigma and perceived stigma (Kosyluk et al., 2016). Personal stigma refers to one’s own beliefs about depression, and perceived stigma refers to one’s beliefs about the attitudes of others (Griffiths et al., 2008). Research on these two types of stigma has reported lower levels of personal stigma comparing to higher perceived stigma (Boerema et al., 2016).

There has been a growing interest in mental health stigma interventions and research in Portugal in the last decade. A European study centered on depression stigma and attitudes towards professional help in the general adult population (Coppens et al., 2013) identified Portugal as the second country (in four) with the highest level of personal and perceived depression stigma, with 31.2% of the participants agreeing with personal stigmatizing affirmations, and 53.8% agreeing with perceived stigma affirmations, measured with the Depression Stigma Scale (Griffiths et al., 2004). In the attitudes towards professional help scale, Portugal ranked in the first place for openness to professional help; however, the value of professional service was not as good, with only 48.2% agreeability. Lower schooling levels, being a male, and not having previous contact with people with mental illness were associated with increased stigma scores (Dietrich et al., 2014).

Particular attention has been made to mental health stigma in health students as health professionals may act as literacy promoters (Xavier et al., 2013). In the past years, research about stigma in health students in Portugal has demonstrated the need for stigma reduction intervention in this group of students (Querido et al., 2016). Anti-stigma-specific education (Gil, 2015; Telles-Correia et al., 2015).

Help-seeking has been a particular problem for first-year university students. Most of the WHO World Mental Health International College Student Initiative report participants reported they would hesitate to seek help if need and attitudinal barriers were identified as the most important (Ebert et al., 2019).

Despite the increasing number of studies addressing mental health stigma and help-seeking (Eisenberg et al., 2009; Musa et al., 2020; Schnyder et al., 2017), most studies neglect to integrate the symptomatology of participants in the analysis.

Also, sampling in young adults has been a problem; usually, participants are health students or professionals (Wei et al., 2015), resulting in a lack of studies contemplating all study areas of the universities.

Lastly, the effects of previous mental healthcare utilization have not been well explored and can be helpful in the study of the relationship between stigma and help-seeking (Eisenberg et al., 2009).

With this study, we aim to (1) examine whether socio-demographic characteristics are related to personal and perceived stigma in students, (2) to understand the effects of the depressive and anxiety symptoms of depression stigma, and (3) examine whether stigmatizing attitudes towards depression is associated with help-seeking behavior in students.

## Methods

### Sample

In February 2019, we contacted all 4493 first-year students from all 14 schools of the University of Porto through institutional email using the SurveyMonkey platform. We invited them to participate in the study. A reminder email was sent a week after the first email.

At the end of the questionnaire, a list of available healthcare responses was available, so students could seek help if they felt it was needed.

### Measures

We asked participants to answer a socio-demographic questionnaire assessing gender, age, place of birth, university course, the Attitudes Toward Seeking Professional Psychological Help (ATSPPH) (Fischer & Turner, 1970), the Patient Health Questionnaire-4 (PHQ-4) (Khubchandani et al., 2016), and the Depression Stigma Scale (DSS) (Griffiths et al., 2004).

The ATSPPH is a ten-item four-point Likert scale, with scores varying from 0 to 30. The scale ranges from “disagree” (score 0) to “agree” (score 3) and comprises two subscales: the first one evaluates the openness to seeking treatment for emotional problems (items 1, 3, 5, 6, and 7), and the second evaluates the value and need on seeking treatment (items 2, 4, 8, 9 and 10, reversed before summed). The maximum score in each subscale is 15, and higher scores indicate better attitudes toward seeking treatment. ATSPPH is the most used scale in mental health help-seeking attitudes, showing excellent psychometric properties in its original form. One of the authors was responsible for the process of translation and adaptation of ATSPPH to the Portuguese population in the context of the OSPI program - Optimizing suicide prevention programs and their implementation in Europe (Coppens et al., 2013; Hegerl et al., 2009; Kohls et al., 2017).

PHQ-4 is a 4-item questionnaire with two items measuring depressive symptoms and two items for anxiety symptoms. Its score varies from 0 to 12; scores 0–4 represent absent symptomatology, 5–8 mild symptomatology, and 9–12 severe symptomatology (Löwe et al., 2010). The PHQ-4 used in the study resulted from the extraction of the corresponding items from the Portuguese version of the PHQ-9 (Ferreira et al., 2018) and GAD-7 (Sousa et al., 2015).

DSS is an 18 five-point Likert scale, with the first nine items measuring personal stigma (one’s own beliefs about depression) and the last nine items measuring perceived stigma (one’s beliefs about the attitudes of others). We converted the total score of both scales as a percentage according to the original score calculation (Griffiths et al., 2004), and the higher the percentage, the higher the stigma. The scale has been translated and validated to the Portuguese population using the OSPI program data (Coppens et al., 2013; Hegerl et al., 2009; Kohls et al., 2017).

### Data analysis

We performed the analysis using SPSS 24 at a 95% confidence level.

We used percentages to describe participation rates, gender distribution, moving away from home, living situation, presence of mental illness in the family, and previous access to mental healthcare. We used counts to describe participant distribution per faculty. Symptomatology stemming from the PHQ-4 scores resulted in three possible severity categories (absent, mild, and severe), and we presented the distribution per category in percentages.

We divided previous access to mental healthcare in Group A, with no previous mental healthcare access, and Group B, with previous access to mental healthcare.

First, we calculated descriptive statistics: frequencies, means, and standard deviations. Then, we performed an analysis of variance to examine gender differences, experience with previous healthcare, symptoms, depression stigma, and help-seeking attitudes using One-Way ANOVA with Tukey HSD’s multiple comparisons test among groups. To test the possible interaction effect between gender and previous mental healthcare groups on depression stigma and help-seeking attitudes, we performed a Two-Way ANOVA.

For normally distributed continuous variables (personal and perceived depression stigma and help-seeking attitudes) and performed T-tests to study differences between the gender, symptomatology groups, and previous access to mental healthcare.

We carried out a Pearson correlation to examine associations between depression stigma and attitudes toward help-seeking.

Lastly, we executed hierarchical linear regressions to statistically control variables to see whether adding variables improves a model’s ability to predict the Depression Stigma and the Help-seeking attitudes scores. Variables included in the models were gender, previous contact with mental illness, mental illness in the family, and symptomatology group.

## Results

### Socio-demographic Characteristics of Participants

Of the 4493 students invited to participate in this study, a total of 1046 (23.3%) accepted to participate. To work with a sample within the late adolescent age limits, we excluded respondents aged 26 years and older, resulting in 969 participants (21.5%). Respondents came from all 14 schools: 197 from Engineering, 164 from Humanities, 111 from Sciences, 82 from Biomedical Sciences, 80 from Economics, 77 from Law, 68 from Psychology and Educational Sciences, 49 from Arts, 44 from Pharmaceutical Sciences, 39 from Medicine, 22 from Nutrition, 16 from Architecture, 13 from Sports, and 7 from Dental Medicine.

About two-thirds of the students were women (64.6%, 626), and 35.4% (343) were men, with a mean age of 18.87 (SD=1.49) ranging between 16 and 25. Approximately 43.9% of the students were away from home, and most of those (46.1%) were living with roommates.

Most participants had never previously sought mental health help care (59.6%), and 35.4% had a family member with a mental illness diagnosis. Of those who had previously sought mental healthcare, anxiety was the main reason (46.4%), followed by depression (25%).

In the PHQ-4, participants showed a mean of 5.65 (SD=3.35), and most of the participants had no symptomatology (51.8%). However, 25.5% showed mild symptomatology and 22.7% severe symptomatology. Only 47.3% of the participants with severe symptomatology had previously sought professional help.

### Personal Depression stigma

Personal depression stigma was very similar between schools, only differing between Arts, Psychology and Educational Sciences and Engineering and Economics, where Arts (M=18.59, SD=9.13) and Psychology and Educational Sciences (M=19.61, SD=9.11) students showed significantly lower mean scores than Engineering (M=26.61, SD=13.57) and Economics (M=26.56, SD=13.07) students.

As shown in Table 1, women had a lower personal stigma level than males. Participants in Group A obtained means of personal depression stigma statistically higher than those in Group B, but these differences turned out to be only significant among women. There was a significant interaction between gender and help-seeking group (F_(968,1)_=4.85, p<0.01).

There was also a significant effect of symptomatology on personal depression stigma, and the mild symptoms groups presented the higher depression stigma score. Tukey HSD’s post-hoc analysis revealed that only the mild symptomatology group differed from the other severity symptoms groups: p<0.01 between absent and mild symptoms and p<0.001 between mild and severe. When comparing depression stigma score differences among help-seeking groups, we verify significance in Group B; however, there was no significant interaction between symptomatology and help-seeking groups (F_(968,2)_=0.92, p=0.40).

Participants with a family member with a mental illness diagnosis had a mean personal stigma score of 22.32 (SD=11.26), a significantly lower result (t_(966)_=-2.24, p<0.05) than those with no family member with mental illness (M=24.17, SD=12.78).

Both gender and the help-seeking group significantly affected personal depression stigma, unlike having a family member with mental illness. Symptomatology was a significant predictor for mild symptoms compared with those with severe symptomatology (Table 2).

The hierarchical multiple regression revealed that at Stage one, gender contributed significantly to the regression model, F_(966,1)_= 48.62, p<0.001, and accounted for 4.7% of the variation in Personal Depression Stigma. Introducing the previous care group variable explained an additional 2.9% variation, and this change in R^2^ was significant, F_(966,1)_= 30.60, p <0.001. Adding family mental illness and symptoms group did not produce an R^2^ significant change. The most important predictor of personal stigma was gender which uniquely explained 4.8% of the variation.

### Perceived depression stigma

Perceived depression mean scores were similar between schools, except for the School of Sports (M=43.60, SD=13.72), which was significantly lower than Sciences (M=60.98, SD=17.71), Biomedical Sciences (M=62.80, SD=17.56), Psychology and Educational Sciences (M=61.84, SD=17.91), Humanities (M=64.78, SD=16.11), Economics (M=64.06, SD=18.83) and Nutrition (M=65.15, SD=18.42).

Gender and help-seeking group differences were statistically significant in the total scores of the perceived depression stigma, with women and help-seeking group B obtaining the highest scores. However, the interaction between gender and the help-seeking group was not significant (F_(968,1)_=0.01, p<0.94).

The severe symptoms group presented the highest perceived stigma score. Tukey HSD’s post-hoc analysis revealed a significant difference between the two groups: no symptoms and severe symptoms (p<0.05). The interaction between the symptoms and help-seeking groups was not significant (F_(968,2)_=0.49, p=0.62).

We obtained non-significant (t_(968)_=1.77, p=0.08) lower scores of perceived stigma in participants with no family member with mental illness (M=60.91, SD=17.31) compared with those with a family member bearing a mental disorder diagnosis (M=63.01, SD=18.49).

Since there were no differences between participants with and without mental illness in the family, we did not include this variable in the hierarchical linear regression.

As can be observed in Table 4, gender and symptomatology scores had significant effect on perceived depression stigma. Despite the significant effects verified, the variance explained in each model is very small (0.6% in Model 1, 0,8% in Model 2, and 1.1% in model 3). Only Model 1 and Model 3 produced a R^2^ significant change: Model 1 F_(967,1)_= 6.41, p <0.05; Model 2 F_(967,1)_= 3.01, p=0.08; Model 3 F_(967,1)_= 4.27, p <0.05.

The most relevant predictor of perceived stigma was gender which explained merely 0.6% of the variation.

### Help-seeking attitudes

Help-seeking attitudes presented a mean of 18.62 (SD=4.76). Women showed significantly higher means (19.20, SD=4.90) than men (17.56, SD=4.32) (t_(966)_ =-5.20, p<0.001). Participants in the help-seeking Group B presented better help-seeking attitudes (20.00, SD=4.54) than those in Group A (17.69, SD=4.69, t_(966)_ =7.66, p<0.001). Differences according to symptomatology were not significant: No symptoms M=18.77, SD=4.93; Mild symptoms M=18.15, SD=4.17; and Severe symptoms M=18.79, SD=4.96 with an ANOVA test of F_(966,2)_= 1.61, p=0.20. We did not find a significant interaction between the global score of the help-seeking attitudes scale and gender, help-seeking group, nor symptomatology group.

Participants with a family member with mental illness presented better help-seeking scores (M=19.22, SD=4.80) than those with no mental illness in the family (M=18.28, SD=4.80), t_(966)_ =2.95, p<0.01.

Personal depression stigma showed a moderate correlation with help-seeking attitudes (r=-0.42, p<0.01). The perceived stigma showed a weaker Pearson correlation with help-seeking attitudes (r=0.10, p<0.01).

The hierarchical multiple regression (table 5) revealed that at Stage one, gender contributed significantly to the regression model, F_(966,1)_= 27.62, p<0.001, and accounted for 2.8% of the variation in the general score of the help-seeking attitudes scale. Introducing the previous care group variable explained an additional 4.9% variation, and this change in R^2^ was significant, F_(966,1)_= 50.80, p <0.001. Adding family mental illness did not produce an R^2^ significant change. Personal depression stigma explained an additional 13.1% of the variance with a significant change in R^2^ (F_(966,1)_= 159.93, p <0.001), and perceived stigma explained an additional 0.01% of the variance. Model 5 total variance explained was 21.6%.

### Openness to seeking treatment for emotional problems

The mean for openness to seeking treatment was 8.82 (SD=2.95), and the mean was higher among women (8.98, SD=3.10) than men (8.53, SD=2.64) with a significant difference: t_(966)_ =-2.67, p<0.05. Participants in help-seeking Group B presented higher mean scores (9.71, SD=2.92) than those in Group A (8.21, SD=2.82), and the differences were statistically significant: t_(966)_ =7.95, p<0.001.

Having a family member with mental illness beneficiated the openness to seeking treatment as the participants in this group obtained a higher mean (9.19, SD=2.88) than those with no mental illness in the family (8.61, SD=2.68), t_(966)_ =2.92, p<0.01.

There were no differences according to symptomatology (F_(966,2)_= 1.29, p=0.28).

Personal depression stigma showed a moderate correlation with openness to seeking treatment (r=-0.30, p<0.01), and the correlation with perceived stigma was not significant.

The hierarchical multiple regression revealed that gender contributed significantly to the regression model at model one, F_(966,1)_= 5.30, p<0.05, but accounted only for 0.05% of the variation. Previous care explained an additional 5.8% variation, and this change in R^2^ was significant, F_(966,1)_= 32.87, p <0.001. Adding the family mental illness group did not produce an R^2^ significant change (F_(966,1)_= 3.67, p=0.06). Personal depression stigma explained an additional 12.7% of the variance with a significant change in R^2^ (F_(966,1)_= 65.81, p <0.001). Introducing perceived stigma in the model also did not result in an R^2^ significant change: F_(966,1)_= 2.07, p=1.51. In this final model, the total variance explained was 12.4%, however only previous care group B (in comparison with group A: β=1.15, CI=0.79, 1.52) and personal stigma (β=-0.06, CI=-0.08, -0.05) had a significant beta regression coefficient.

### Value and need of seeking treatment

Participants showed a mean of 9.81 (SD=2.56) in the value and need of seeking treatment. Woman presented significant higher means (10.23, SD=2.50) than men (9.03, SD=2.49), t_(966)_ =-7.01, p<0.001. Participants who have previously sought help (Group B) obtained better mean scores (10.29, SD=2.38) than participants in Group A (9.47, SD=2.63), with differences statistically significant: t_(966)_ =4.97, p<0.001. Participants with mental illness in the family presented better mean scores (10.03, SD=2.48) than participants with no family member with mental illness (9.67, SD=2.59), and these differences were statistically significant: t_(966)_ =2.10, p<0.05.

Symptomatology did not impact mean scores for value and need of seeking treatment (F_(966,2)_= 1.83, p=0.16).

Personal depression stigma showed a moderate correlation with openness to seeking treatment (r=-0.45, p<0.001), and the correlation with perceived also showed a significant correlation, yet weaker (r=0.10, p<0.01).

Gender accounted for 5% of the variation as the first variable included in the hierarchical multiple regression, with a significant contribution for the R^2^ change: F_(966,1)_= 51.37, p<0.001. The previous care group variable explained an additional 1.8% variation, and the change produced in R^2^ was also significant, F_(966,1)_= 17.79, p <0.001. Family mental illness group did not change the R^2^ (F_(966,1)_= 1.58, p=0.21. Personal depression stigma explained an additional 15.3% of the variance with a significant change in R^2^ (F_(966,1)_= 189.38, p <0.001), and perceived stigma explained an additional lower amount of variance (0.1%) yet significant in the R^2^ change: F_(966,1)_= 8.94, p<0.01. Model 5 explained a total variance of 22.9%. The previous help group and family with mental illness group did not have a significant beta regression coefficient.

## Discussion

We contacted students to participate in this study via their institutional email addresses. Since they do not access their institutional email address very frequently, response rates were relatively low. Thus, the response rate was lower than the mean in similar studies in the general population (Burgard et al., 2019). Another explanation may be the recent increase in online surveys. Nevertheless, there was a comparable response rate within each school, except for Economics, with a much higher response rate than the mean, and Sports and Architecture, with a response rate lower than 10%.

The gender distribution is comparable to the population: in the school year of 2018-2019, the University of Porto’s records show that 59% of the students were women and 41% men.

Our sample presented lower stigma scores than the previous study in the general population (Coppens et al., 2013), which may be because our sample presents a much lower mean age, and depression stigma is associated with older age (Coppens et al., 2013; Dietrich et al., 2014).

As expected, personal stigma showed significantly lower means than perceived stigma, and women showed lower levels of personal stigma and higher levels of perceived stigma (Boerema et al., 2016). Some authors argue that may be due to masculine norms (Chatmon, 2020), a set of social norms, and expected behavior associated with men (Milner et al., 2018).

The previous use of mental health care had a significant effect on the personal stigma of women, but not men, which may also be because men have a more stigmatized perspective on mental illness. Thus, access to mental health care may not have the same stigma-reducing effect on men and women.

For perceived stigma, the difference becomes insignificant when analyzing the results by gender. This result agrees with previous studies, where the effects of the previous contact with depression are more evident for personal stigma than perceived stigma (Busby Grant et al., 2016; Griffiths et al., 2008).

Depressive symptoms have a mitigating effect on personal stigma for those who have previously sought help, which may be due to the personal stigma reduction effects of mental health interventions.

Participants with a family member with mental illness, and thus having indirect experience with mental illness, showed lower personal depression stigma means; however, its effect on personal stigma was not significant.

Participants with milder symptoms showed more personal stigma and lower perceived stigma than those with severe symptomatology, resulting from a process of self-stigmatization and self-denial of a possible need for help, especially in the case of personal stigma. This particularity may be an essential element to consider when addressing personal stigma.

Symptomatology did not impact help-seeking attitudes, suggesting the importance of mental health literacy promotion to improve the recognized value of health care when needed.

The effect of gender is more substantial in value and need of seeking treatment than on the openness to seek treatment. While women see more value and need in the treatment-seeking, there are no significant differences when other effects are considered, such as personal stigma.

Past help-seeking had a more significant effect on the openness to seek treatment than on the value and need of seeking treatment, which may be interesting to research further as previous help-care experiences are an essential tool for literacy and help-seeking behaviors promotion.

Demonstration of a more substantial correlation of depression personal stigma with help-seeking attitudes than gender or previous contact with mental healthcare services confirms the importance of stigma interventions in promoting help-seeking behavior (Gulliver et al., 2010; Thornicroft et al., 2017). Personal stigma concerning depression was the variable with the most significant magnitude effect on help-seeking attitudes, in line with the strong effect of attitudinal barriers in help-seeking (Ebert et al., 2019).

This result reinforces the importance of reducing stigma as a strategy to promote help-seeking behaviors.

One of the study’s main limitations is the sample’s representativeness since the high rate of previous mental healthcare service usage, and the distributions of the PHQ-4 scores suggest some degree of selection bias. The second limitation is the lack of data on essential elements of stigma, namely the time and duration of mental health access care. Further studies should consider these elements to understand better the effects of previous mental healthcare usage on stigma and help-seeking attitudes.

Despite the possible and inevitable sample bias in online studies, our sample compares well to the University of Porto population, both in distribution by the school as distribution by gender, strengthening the generalizability of the results.

Due to our sample’s low age variance, we only considered first-year students below 25 years old – age was not included as a study variable. Future research directed to the understanding of the relationship between age and depression stigma will address this gap. As this particular sample may be followed, the possible effect of stigma reduction as a result of the universities experiences and learnings, as reported on Portuguese health students (Querido et al., 2016).

In conclusion, intensifying mental health promotion and easing access to mental healthcare will increase the probability of successful early intervention on college student’s depression and anxiety disorders. More research will be needed to understand the different effects of mental health services on stigma reduction among men and women.

These findings also reinforce the importance of stigma reduction interventions as a tool to promote help-seeking behaviors. Future research on the actual effect of stigma reduction on healthcare use is therefore essential.

## Data Availability

All data refered in the manuscript may be available upon request to the corresponding author

## References

Alonso, J., Mortier, P., Auerbach, R. P., Bruffaerts, R., Vilagut, G., Cuijpers, P., Demyttenaere, K., Ebert, D. D., Ennis, E., Gutierrez-Garcia, R. A., Green, J. G., Hasking, P., Lochner, C., Nock, M. K., Pinder-Amaker, S., Sampson, N. A., Zaslavsky, A. M., Kessler, R. C., & Collaborators, W. W.-I. (2018, Sep). Severe role impairment associated with mental disorders: Results of the WHO World Mental Health Surveys International College Student Project. Depress Anxiety, 35(9), 802–814. https://doi.org/10.1002/da.22778

Arnett, J. J. (2000). Emerging adulthood: A theory of development from the late teens through the twenties. American Psychologist, 55(5), 469–480. https://doi.org/10.1037/0003-066x.55.5.469

Auerbach, R. P., Mortier, P., Bruffaerts, R., Alonso, J., Benjet, C., Cuijpers, P., Demyttenaere, K., Ebert, D. D., Green, J. G., Hasking, P., Murray, E., Nock, M. K., Pinder-Amaker, S., Sampson, N. A., Stein, D. J., Vilagut, G., Zaslavsky, A. M., Kessler, R. C., & Collaborators, W. W.-I. (2018, Oct). WHO World Mental Health Surveys International College Student Project: Prevalence and distribution of mental disorders. J Abnorm Psychol, 127(7), 623–638. https://doi.org/10.1037/abn0000362

Boerema, A. M., Zoonen, K., Cuijpers, P., Holtmaat, C. J., Mokkink, L. B., Griffiths, K. M., & Kleiboer, A. M. (2016). Psychometric Properties of the Dutch Depression Stigma Scale (DSS) and Associations with Personal and Perceived Stigma in a Depressed and Community Sample. PLoS One, 11(8), e0160740. https://doi.org/10.1371/journal.pone.0160740

Burgard, T., Kasten, N., & Bosnjak, M. (2019). Participation in online surveys in psychology. A meta-analysis. Research Synthesis 2019 conference, Dubrovnik, Croatia.

Busby Grant, J., Bruce, C. P., & Batterham, P. J. (2016, Jun). Predictors of personal, perceived and self-stigma towards anxiety and depression. Epidemiol Psychiatr Sci, 25(3), 247–254. https://doi.org/10.1017/S2045796015000220

Chatmon, B. N. (2020, Jul-Aug). Males and Mental Health Stigma. Am J Mens Health, 14(4), 1557988320949322. https://doi.org/10.1177/1557988320949322

Coppens, E., Van Audenhove, C., Scheerder, G., Arensman, E., Coffey, C., Costa, S., Koburger, N., Gottlebe, K., Gusmao, R., O’Connor, R., Postuvan, V., Sarchiapone, M., Sisask, M., Szekely, A., van der Feltz-Cornelis, C., & Hegerl, U. (2013, Sep 5). Public attitudes toward depression and help-seeking in four European countries baseline survey prior to the OSPI-Europe intervention. J Affect Disord, 150(2), 320–329. https://doi.org/10.1016/j.jad.2013.04.013

Corrigan, P. W. (2006). Mental Health Stigma as Social Attribution: Implications for Research Methods and Attitude Change. Clinical Psychology: Science & Practice, 7(1), 48–67.

Dietrich, S., Mergl, R., & Rummel-Kluge, C. (2014, Dec 15). Personal and perceived stigmatization of depression: a comparison of data from the general population, participants of a depression congress and job placement officers in Germany. Psychiatry Res, 220(1-2), 598–603. https://doi.org/10.1016/j.psychres.2014.06.044

Ebert, D. D., Mortier, P., Kaehlke, F., Bruffaerts, R., Baumeister, H., Auerbach, R. P., Alonso, J., Vilagut, G., Martínez, K. I., Lochner, C., Cuijpers, P., Kuechler, A. M., Green, J., Hasking, P., Lapsley, C., Sampson, N. A., & Kessler, R. C. (2019, Jun). Barriers of mental health treatment utilization among first-year college students: First cross-national results from the WHO World Mental Health International College Student Initiative. Int J Methods Psychiatr Res, 28(2), e1782. https://doi.org/10.1002/mpr.1782

Eisenberg, D., Downs, M. F., Golberstein, E., & Zivin, K. (2009, Oct). Stigma and help seeking for mental health among college students. Med Care Res Rev, 66(5), 522–541. https://doi.org/10.1177/1077558709335173

Ferreira, T., Sousa, M., & Salgado, J. (2018). Brief assessment of depression: Psychometric properties of the Portuguese version of the Patient Health Questionnaire (PHQ-9). The Psychologist: Practice & Research Journal, 1(2), 1–12.

Fischer, E. H., & Turner, J. L. (1970). Development and Research Utility of an Attitude Scale. Journal of Consulting and Clinical Psychology, 35(1), 70–90.

Gil, I. (2015). Estigma e doença mental. In Cadernos de Psiquiatria social e cultural (pp. 59–76). https://doi.org/10.14195/978-989-26-0968-3_3

Griffiths, K. M., Christensen, H., & Jorm, A. F. (2008, Apr 18). Predictors of depression stigma. BMC Psychiatry, 8, 25. https://doi.org/10.1186/1471-244X-8-25

Griffiths, K. M., Christensen, H., Jorm, A. F., Evans, K., & Groves, C. (2004). Effect of Web-Based Depression Literacy and Cognitive-Behavioural Therapy Interventions on Stigmatising Attitudes to Depression: Randomised Controlled Trial. The British Journal of Psychiatry (185), 342–349. https://doi.org/10.1192/bjp.185.4.342

Gulliver, A., Griffiths, K. M., & Christensen, H. (2010). Perceived barriers and facilitators to mental health help-seeking in young people: a systematic review. BMC Psychiatry, 10(113).

Hegerl, U., Wittenburg, L., Arensman, E., Van Audenhove, C., Coyne, J. C., McDaid, D., van der Feltz-Cornelis, C. M., Gusmao, R., Kopp, M., Maxwell, M., Meise, U., Roskar, S., Sarchiapone, M., Schmidtke, A., Varnik, A., & Bramesfeld, A. (2009, Nov 23). Optimizing suicide prevention programs and their implementation in Europe (OSPI Europe): an evidence-based multi-level approach. BMC Public Health, 9, 428. https://doi.org/10.1186/1471-2458-9-428

Khalema, N. E., & Shankar, J. (2014). Perspectives on Employment Integration, Mental Illness and Disability, and Workplace Health. Advances in Public Health, 2014.

Khubchandani, J., Brey, R., Kotecki, J., Kleinfelder, J., & Anderson, J. (2016). The Psychometric Properties of PHQ-4 Depression and Anxiety Screening Scale Among College Students. Archives of Psychiatric Nursing, 30(4), 457–462.

Kohls, E., Coppens, E., Hug, J., Wittevrongel, E., Van Audenhove, C., Koburger, N., Arensman, E., Szekely, A., Gusmao, R., & Hegerl, U. (2017, Aug 1). Public attitudes toward depression and help-seeking: Impact of the OSPI-Europe depression awareness campaign in four European regions. J Affect Disord, 217, 252–259. https://doi.org/10.1016/j.jad.2017.04.006

Kosyluk, K. A., Al-Khouja, M., Bink, A., Buchholz, B., Ellefson, S., Fokuo, K., Goldberg, D., Kraus, D., Leon, A., Michaels, P., Powell, K., Schmidt, A., & Corrigan, P. W. (2016, Sep). Challenging the Stigma of Mental Illness Among College Students. J Adolesc Health, 59(3), 325–331. https://doi.org/10.1016/j.jadohealth.2016.05.005

Löwe, B., Wahl, I., Rose, M., Spitzer, C., Glaesmer, H., Wingenfeld, K., Schneider, A., & Brähler, E. (2010). A 4-item measure of depression and anxiety: Validation and standardization of the Patient Health Questionnaire-4 (PHQ-4) in the general population. Journal of Affective Disorders, 122, 86–95.

McNair, B. G., Highet, N. J., Hickie, I. B., & Davenport, T. A. (2002). Exploring the Perspectives of People Whose Lives Have Been Affected by Depression. The Medical Journal of Australia, 196(10), 69–76.

Milner, A., Kavanagh, A., King, T., & Currier, D. (2018, Jul). The Influence of Masculine Norms and Occupational Factors on Mental Health: Evidence From the Baseline of the Australian Longitudinal Study on Male Health. Am J Mens Health, 12(4), 696–705. https://doi.org/10.1177/1557988317752607

Moritz, D., & Roberts, J. E. (2016, 2016/10/01/). Individual Differences in Propensity to Socially Reject Depressed Others. Personality and Individual Differences, 101, 500. https://doi.org/10.1016/j.paid.2016.05.233

Musa, A., Ashraf, J., Tsai, F. J., Abolmagd, S., Liu, C., Hussain, H., Voslarova, E., Khalil, M. A., Wolitzky-Taylor, K. B., Lee, D., Sugar, J., Pendi, K., Lee, J., Abdelmaksoud, R., Adel, N., & Baron, D. (2020, Nov). Depression Severity and Depression Stigma Among Students: A Survey of Universities in Five Countries. J Nerv Ment Dis, 208(11), 884–889. https://doi.org/10.1097/NMD.0000000000001226

Querido, A., Tomás, C., & Carvalho, D. (2016). O estigma face à doença mental nos estudantes de saúde. Revista Portuguesa de Enfermagem de Saúde Mental(Spe. 3). https://doi.org/10.19131/rpesm.0120

Sawyer, S. M., Azzopardi, P. S., Wickremarathne, D., & Patton, G. C. (2018). The age of adolescence. The Lancet Child & Adolescent Health, 2(3), 223–228. https://doi.org/10.1016/s2352-4642(18)30022-1

Schnyder, N., Panczak, R., Groth, N., & Schultze-Lutter, F. (2017, Apr). Association between mental health-related stigma and active help-seeking: systematic review and meta-analysis. Br J Psychiatry, 210(4), 261–268. https://doi.org/10.1192/bjp.bp.116.189464

Sousa, T. V., Viveiros, V., Chai, M. V., Vicente, F. L., Jesus, G., Carnot, M. J., Gordo, A. C., & Ferreira, P. L. (2015, Apr 25). Reliability and validity of the Portuguese version of the Generalized Anxiety Disorder (GAD-7) scale. Health Qual Life Outcomes, 13, 50. https://doi.org/10.1186/s12955-015-0244-2

Telles-Correia, D., Marques, J. G., Gramaça, J., & Sampaio, D. (2015). Stigma and Attitudes towards Psychiatric Patients in Portuguese Medical Students. Acta Médica Portuguesa, 28(6), 715–719.

Thornicroft, G., Chatterji, S., Evans-Lacko, S., Gruber, M., Sampson, N., Aguilar-Gaxiola, S., Al-Hamzawi, A., Alonso, J., Andrade, L., Borges, G., Bruffaerts, R., Bunting, B., de Almeida, J. M., Florescu, S., de Girolamo, G., Gureje, O., Haro, J. M., He, Y., Hinkov, H., Karam, E., Kawakami, N., Lee, S., Navarro-Mateu, F., Piazza, M., Posada-Villa, J., de Galvis, Y. T., & Kessler, R. C. (2017, Feb). Undertreatment of people with major depressive disorder in 21 countries. Br J Psychiatry, 210(2), 119–124. https://doi.org/10.1192/bjp.bp.116.188078

Uehara, T., Takeuchi, K., Kubota, F., Oshima, K., & Ishikawa, O. (2010). Annual transition of major depressive episode in university students using a structured self-rating questionnaire. Asia-Pacific Psychiatry, 2(2), 99–104. https://doi.org/10.1111/j.1758-5872.2010.00063.x

Wei, Y., McGrath, P. J., Hayden, J., & Kutcher, S. (2015, Nov 17). Mental health literacy measures evaluating knowledge, attitudes and help-seeking: a scoping review. BMC Psychiatry, 15, 291. https://doi.org/10.1186/s12888-015-0681-9

World Health Organization. (2001). The WHO World Health Report 2001: New understanding - New hope. W. L. Cataloguing.

World Health Organization. (2013). Mental Health Action Plan 2013-2020.

World Health Organization. (2017). Depression and Other Common Mental Disorders: Global Health Estimates.

Xavier, S., Klut, C., Neto, A., Ponte, G. d., & Melo, J. C. (2013). O Estigma da Doença Mental: Que Caminho Percorremos? Revista do Serviço de Psiquiatria do Hospital Prof. Doutor Fernando Fonseca, EPE, 11(2), 10–21.

