## Supplementary file for "The association between stigmatizing attitudes towards depression and help seeking attitudes in college students"

Table 1: Personal and perceived depression stigma mean scores per school

|  |  | **n** | **Personal Depression Stigma**  **M (SD)** | **Perceived Depression Stigma**  **M (SD)** |
| --- | --- | --- | --- | --- |
| Engineering | | 197 | 26.61 (13.57) | 59.75 (26.95) |
| Humanities | | 164 | 24.07 (12.06) | 64.78 (16.11) |
| Sciences | | 111 | 23.12 (12.01) | 60.98 (17.71) |
| Biomedical Sciences | | 82 | 21.81 (10.03) | 62.80 (17.56) |
| Economics | | 80 | 26.56 (13.07) | 64.06 (18.83) |
| Law | | 77 | 22.72 (15.90) | 60.35 (20.81) |
| Psychology and Educational Sciences | | 68 | 19.61 (9.11) | 61.84 (17.91) |
| Arts | | 49 | 18.59 (9.13) | 60.43 (17.94) |
| Pharmaceutical Sciences | | 44 | 24.05 (12.23) | 59.78 (17.63) |
| Medicine | | 39 | 22.22 (9.32) | 60.68 (13.31) |
| Nutrition | | 22 | 21.59 (11.78) | 65.15 (18.42) |
| Architecture | | 16 | 26.21 (10.92) | 62.67 (13.72) |
| Sports | | 13 | 26.07 (10.67) | 43.60 (13.72) |
| Dental Medicine | | 7 | 23.02 (12.39) | 54.76 (16.64) |

Table 2: Personal and perceived depression stigma Tukey HSD's post-hoc significant differences between schools.

|  | Schools | | M | SE | p | CI |
| --- | --- | --- | --- | --- | --- | --- |
| Personal Depression Stigma | Arts | Engineering | -8.01 | 1.95 | <0.01 | -14.58, -1.45 |
|  |  | Economics | -7.97 | 2.21 | <0.05 | -15.43, -0.51 |
|  | Psychology and Educational Sciences | Engineering | -6.99 | 1.72 | <0.01 | -12.78, -1.22 |
|  |  | Economics | -6.95 | 2.02 | <0.05 | -13.74, -0.17 |
| Perceived Depression Stigma | Sports | Sciences | -17.40 | 5.16 | <0.05 | -34.75, -0.04 |
|  |  | Biomedical Sciences | -19.21 | 5.26 | <0.05 | -36.89, -1.54 |
|  |  | Psychology and Educational Sciences | -18.26 | 5.33 | <0.05 | -36.17, -0.33 |
|  |  | Humanities | -21.20 | 5.07 | <0.01 | -38.26, -4.14 |
|  |  | Economics | -20.47 | 5.27 | <0.01 | -38.18, 2.77 |
|  |  | Nutrition | -21.56 | 6.16 | <0.05 | -42.27, -0.84 |

Table 3: Effects of gender, previous help group, family mental illness, personal depression stigma and perceived depression stigma on Openness to seeking treatment for emotional problems

|  | **β** | **95% CI** | **t** | **p** |
| --- | --- | --- | --- | --- |
|  | **Model 1** |  |  |  |
| Women | Ref. |  |  |  |
| Men | **-0.45** | **-0.84, -0.06** | **2.30** | **<0.05** |
|  | **Model 2** |  |  |  |
| Women | Ref. |  |  |  |
| Men | -0.28 | -0.07, 0.10 | 1.48 | 0.14 |
| Previous help – Group A | Ref. |  |  |  |
| Previous help – Group B | **1.47** | **1.10, 1.84** | **-7.75** | **<0.001** |
|  | **Model 3** |  |  |  |
| Women | Ref. |  |  |  |
| Men | -0.27 | -0.65, 0.11 | 1.40 | 0.16 |
| Previous help – Group A | Ref. |  |  |  |
| Previous help – Group B | **1.42** | **1.05, 1.79** | **-7.46** | **<0.001** |
| Family with mental illness - No | Ref. |  |  |  |
| Family with mental illness - Yes | 0.37 | -0.01, 0.75 | -1.92 | 0.06 |
|  | **Model 4** |  |  |  |
| Women | Ref. |  |  |  |
| Men | 0.04 | -0.34, 0.41 | -0.21 | 0.83 |
| Previous help – Group A | Ref. |  |  |  |
| Previous help – Group B | **1.17** | **0.80, 1.53** | **-6.23** | **<0.001** |
| Family with mental illness – No | Ref. |  |  |  |
| Family with mental illness – Yes | 0.30 | -0.07, 0.67 | -1.58 | 0.11 |
| Personal Depression Stigma | **-0.06** | **-0.08, -0.05** | **-8.11** | **<0.001** |
|  | **Model 5** |  |  |  |
| Women | Ref. |  |  |  |
| Men | -0.62 | -0.21, 0.44 | -0.32 | 0.75 |
| Previous help – Group A | Ref. |  |  |  |
| Previous help – Group B | **1.15** | **0.79, 1.52** | **-6.15** | **<0.001** |
| Family with mental illness - No | Ref. |  |  |  |
| Family with mental illness - Yes | 0.29 | -0.08, 0.65 | -1.52 | 0.13 |
| Personal Depression Stigma | **-0.06** | **-0.08, -0.05** | **-8.16** | **<0.001** |
| Perceived Depression Stigma | 0.01 | -0.01, 0.02 | 1.44 | 0.15 |

β=beta regression coefficients, Ref.=Reference category

* Model 1= gender; Model 2: Model 1 plus help-seeking group; Model 3: Model 2 plus family mental illness; Model 4: Model 3 plus Personal Depression Stigma; Model 5: Model 4 plus Perceived Depression Stigma.

Significant results are in bold.

Table 4: Effects of gender, previous help group, family mental illness, personal depression stigma and perceived depression stigma on Value and need of seeking treatment

|  | **β** | **95% CI** | **t** | **p** |
| --- | --- | --- | --- | --- |
|  | **Model 1** |  |  |  |
| Women | Ref. |  |  |  |
| Men | **-1.20** | **-1.53, -0.87** | **7.17** | **<0.001** |
|  | **Model 2** |  |  |  |
| Women | Ref. |  |  |  |
| Men | **-1.12** | **-1.45, -0.79** | **6.70** | **<0.001** |
| Previous help – Group A | Ref. |  |  |  |
| Previous help – Group B | **0.67** | **0.37, 1.00** | **-4.03** | **<0.001** |
|  | **Model 3** |  |  |  |
| Women | Ref. |  |  |  |
| Men | **-1.11** | **-1.44, -0.79** | **6.65** | **<0.001** |
| Previous help – Group A | Ref. |  |  |  |
| Previous help – Group B | **0.66** | **0.34, 0.99** | **-4.02** | **<0.001** |
| Family with mental illness - No | Ref. |  |  |  |
| Family with mental illness - Yes | 0.21 | -0.12, 0.54 | -1.26 | 0.21 |
|  | **Model 4** |  |  |  |
| Women | Ref. |  |  |  |
| Men | **-0.68** | **-0.99, -0.38** | **4.37** | **<0.001** |
| Previous help – Group A | Ref. |  |  |  |
| Previous help – Group B | **0.31** | **0.01, 0.61** | **-2.02** | **<0.05** |
| Family with mental illness – No | Ref. |  |  |  |
| Family with mental illness – Yes | 0.11 | -0.19, 0.41 | -0.70 | 0.48 |
| Personal Depression Stigma | **-0.08** | **-0.10, -0.07** | **-13.76** | **<0.001** |
|  | **Model 5** |  |  |  |
| Women | Ref. |  |  |  |
| Men | **-0.65** | **-0.95, -0.34** | **4.14** | **<0.001** |
| Previous help – Group A | Ref. |  |  |  |
| Previous help – Group B | 0.28 | -0.01, 0.58 | -1.87 | 0.06 |
| Family with mental illness - No | Ref. |  |  |  |
| Family with mental illness - Yes | 0.09 | -0.21, 0.37 | -0.56 | 0.57 |
| Personal Depression Stigma | **-0.09** | **-0.10, -0.07** | **-13.91** | **<0.001** |
| Perceived Depression Stigma | **0.01** | **0.01, 0.02** | **2.99** | **<0.01** |

β=beta regression coefficients, Ref.=Reference category

* Model 1= gender; Model 2: Model 1 plus help-seeking group; Model 3: Model 2 plus family mental illness; Model 4: Model 3 plus Personal Depression Stigma; Model 5: Model 4 plus Perceived Depression Stigma.

Significant results are in bold.
